# ‘It just adds that other dimension to your decision making’: findings from a UK study with adult safeguarding practitioners of a pilot forensic service to support adult safeguarding

**DOI:** 10.64898/2026.10.23.26365439

**Authors:** Caroline White, Dave Marsland

## Abstract

**Purpose:** This study explored the contribution of a Forensic Service for adult safeguarding introduced in one geographic area. This service introduced the role of Forensic Medical Examiner (FME) to examine and interpret injuries when there are concerns that these may have non-accidental causes. This paper explores the perspectives of adult safeguarding Social Workers and the FME about their experiences of the service and how it supported their decision-making and safeguarding practice.

**Methodology:** Semi-structured interviews were conducted with the FME and Social Workers from safeguarding adults teams. The data was analysed using thematic analysis.

**Findings:** Four themes were generated through the analysis. These highlighted the challenges for Social Workers and other practitioners in identifying whether a non-accidental injury has occurred; the role of forensic information in enhancing knowledge and understanding; the contribution of forensic information to safeguarding decision making and activity; Social Worker and FME experiences of working together within the new service.

**Originality:** The Forensic Service is, to the best of our understanding, the first service of its kind within the United Kingdom. It appears to represent a unique response to the challenges of interpreting injuries and determining when these may be non-accidental, and attributable to physical abuse or neglect.

## Introduction

The abuse of adults with care and support needs is an ongoing problem internationally, often causing significant impacts to those abused (Rowsell *et al*; Yunus *et al*., 2019). However, detecting and bringing abuse to light is beset with challenges. While it is important not to overlook the abilities of those who have been abused to speak out about their experiences, there are multiple barriers to disclosure. These include individuals fearing the consequences of reporting, concerns about loss of relationships, worrying they will not be taken seriously or minimising the seriousness of what they are experiencing; communication difficulties, cognitive impairments or lack of capacity also impede disclosure (Gironda *et al*., 2016; Dow *et al*., 2020; Walsh *et al*., 2024). Accordingly, approaches to detecting abuse cannot rely solely on disclosure, and Prichard-Jones *et al* (2022, p.133) have attested that adult safeguarding is:

> *An area of practice that depends as much on what practitioners are seeing around the person potentially being abused or neglected, as on the conversations they have with that person*.

Abuse is frequently hidden; practitioners seldom directly witness acts of abuse and neglect, and are often reliant on interpreting more ambiguous signs which may indicate a range of causes, including but not restricted to abuse (Marsland *et al*., 2015; White and Alton, 2024). Identifying and responding to possible abuse and neglect is therefore fraught with difficulties, characterised by complexity and uncertainty, and may be underpinned by *contested accounts, vague or retracted disclosures, deception and inconclusive evidence* (Thacker *et al*., 2019, pp.258-259).

Physical signs and injuries may arise through non-accidental injury (NAI), which may signify abuse and neglect. Determining whether injuries such as bruising or fractures have arisen accidentally or indicate abuse may be challenging (Wiglesworth *et al*., 2009; Gironda et *al*., 2016; van Houten *et al*., 2021). However, there are emerging understandings of patterns of injuries associated with non-accidental causes, which can assist in identifying physical abuse and neglect (Mosqueda et *al*., 2005: Wiglesworth *et al*., 2009; van Houten *et al*., 2022, Rosen *et al*., 2020: Yonashiro-Cho *et al*., 2021) by skilled and trained practitioners. Service developments have been documented in some areas of North America. These employ multi-agency teams to respond to adult abuse and include harnessing the forensic skills of clinicians and knowledge of injury patterns to support examination and assessment when abuse is suspected, (Mosqueda *et al*., 2004; Wiglesworth *et al*., 2006; Rosen *et al*., 2022; Bloemen *et al*., 2024). However, we are aware of no equivalent services in the UK, and overall this appears a neglected area internationally, highlighting the significance of the learning from this service innovation.

In England, the Care Act (2014) mandates that local authorities are the lead agency for making safeguarding enquiries (although other agencies may undertake such work), with Social Workers therefore having a key role in responding to suspected abuse (Department of Health and Social Care, 2025). Social Workers within children’s services have access to Child Protection Medical Examinations to examine and document injuries and determine potential causes (Garstang *et al*., 2021). However, there is no comparable service which provides expertise in interpreting injuries in adults. In most instances in which there is a concern about injury patterns there is no recourse to an expert examination or opinion, although the need for this has been highlighted as an important skill for safeguarding practitioners (Royal College of Nursing, 2018). A Safeguarding Adults Review (SAR; a review mandated by the Care Act 2014 in the event of serious abuse/neglect, including death, of an adult) noted the significance of this lack of access to forensic expertise:

> *Doctors may well be concerned regarding patterns of bruising, but without a forensic medical training and/or background, would be unlikely [to] be able to give the definitive forensic medical opinion….required and is a gap in the system to safeguard adults (Rees, 2022, p,9)*.

This SAR and the systematic gap highlighted was instrumental in the development of a pilot Forensic Service for Adult Safeguarding within one geographic area in England. This service represents a partnership between an Integrated Care Board (ICB, NHS body responsible for planning and commissioning health services) and two local authorities. At the centre is a general practitioner (GP; family doctor) who is also a Named Doctor for Safeguarding Adults, who has undertaken training in forensic examination, and whose role is that of the Forensic Medical Examiner (FME) for the service. They are employed to support the service on a part time-basis. They work closely with:

- Adult safeguarding Social Workers within the two local authorities. Their role includes working with the FME in respect of referral, information gathering, supporting examination visits
- The Police, who are involved when a crime is suspected
- Health practitioners, who may support investigations.

### The pilot Forensic Service

The service was launched in 2022, following preparatory work which included training for the FME; steering group formation; policy development (Alton et al., 2023), with initial funding from the Clinical Commissioning Groups (former NHS bodies) within the local area. Its aims were to provide an examination service for adults at risk of harm, where there were concerns they may have sustained NAI as a consequence of physical abuse or neglect. The Forensic Service does not provide treatment, but undertakes examination, documentation and interpretation of injuries, and contributes to safeguarding enquiries conducted under Section 42 of the Care Act 2014, and subsequent safeguarding planning.

Referrals to the Forensic Service are made by Social Workers in the Adult Safeguarding Teams within the two local authorities if concerns are raised by referrers or become apparent during triage of incoming safeguarding referrals. Following referral there is usually a discussion between the FME and a safeguarding adults Social Worker; Police may also be involved if a crime is suspected. If a forensic examination is indicated, a joint visit is undertaken by the FME and a Social Worker. The FME conducts the examination (usually accompanied by someone who knows the person, can facilitate communication and help them feel more comfortable), while the Social Worker gathers further information, for example, through examining care plans and records which may provide information indicating potential causes of injuries. Examinations usually take place in the person’s own home or place where they are receiving care, recognising that, for many, travelling to another location for examination would be impractical and potentially disorienting. Consent is taken prior to examination, with Best Interest processes followed if individuals lack capacity to consent to the examination, in line with the Mental Capacity Act. Although it was expected that examinations would occur in-person, photographs are also used, for example, if there has been a delay in the referral meaning that injuries have faded and cannot be documented through a physical examination. Safeguarding Social Workers may also share photographs obtained as part of a referral to seek advice about whether an examination or further investigation appears warranted, with a photography policy developed in respect of taking and sharing photographs of injuries.

Data collected by the FME from May 2022 – March 2023 provides information in respect of the characteristics and needs of those examined, although missing data was noted (White and Marsland, 2023). This data highlighted that:

- Almost half of those examined (49%) were aged 80 years or over.
- 64% were identified as a frail older person, a person living with dementia or both, a further 14.5% were adults with learning disabilities.
- 74.5% lived in a care home, relatively few (approximately 7%) lived in their own homes, either alone or with others.
- The majority of those examined (69%) were assessed as lacking capacity to consent to the examination.

## Methods

The data reported in this paper were collected as part of a wider evaluation of the Forensic Service (White and Marsland, 2023) which explored the early development and delivery of the pilot service, including the work to develop training, policies and procedures, the conduct of examinations and the multi-agency work undertaken. While the work of a small number of services internationally which incorporate forensic skills have been documented, there has been little focus on how medical and Social Work expertise are brought together to respond to suspected abuse. This paper seeks to address this gap in the evidence base, and brings together data from interviews with the FME who conducted the examinations, and adult safeguarding Social Workers. It seeks to address the following questions:

> *How does the introduction of a Forensic Examination Service enable health and Social Work practitioners to support adults who may have experienced non-accidental injury?*
>
> *How do Social Workers and Forensic Medical Examiners work together to capture and use forensic and other information to support adult safeguarding decisions?*

### Participant recruitment

Emails were sent to Social Workers within the Safeguarding Adults teams, inviting them to participate in the research. Those expressing interest were forwarded a written Participant Information Sheet, which was also discussed immediately prior to interview. All were asked to sign a consent form before interview.

### Data collection

All participants took part in in-depth interviews, guided by a semi-structured interview schedule. Interviews were conducted online or in-person, recorded and transcribed verbatim between August 2022 and January 2023.

### Data analysis

The data were analysed using the six steps of thematic analysis outlined by Braun and Clarke (2006). These include: familiarisation; generating initial codes; searching for, reviewing and naming themes; reporting findings. Interview transcripts were read closely for familiarisation, and initial codes developed inductively from the data. These initial codes were grouped into four overall themes, which were named to reflect the key meanings within each theme. NVivo14 was used to manage data and support the coding process.

### Ethical issues

The study received ethical approval from the University of Hull Faculty of Health Sciences Research Ethics Committee. All participants gave consent for the use of quotes, which were anonymised with gender neutral terms added to further preserve anonymity for participants and for those for whom there have been safeguarding concerns.

## Findings

### Participants

In addition to the FME who was interviewed three times during the research, six adult safeguarding Social Workers participated (three from each local authority). All worked within the Adult Safeguarding Teams; five were Social Workers (including three Senior Social Workers) and one a manager.

From the analysis of the transcripts, 4 themes were identified:

- Uncertainties in recognising and identifying non-accidental injury among Social Workers and other practitioners
- The role of forensic expertise in enhancing knowledge and understanding of non-accidental injury
- The role of forensic information in supporting safeguarding decision making and activity
- Working together within the new service partnership

### Theme One – Uncertainties in recognising and identifying non-accidental injury among Social Workers and other practitioners: ‘It’s really hard, isn’t it, to say for sure’

Social Workers described feelings of uncertainty, concern, *uneasy feelings (Social Worker 3)*, and suspicion in respect of things they see or information provided on referral:

> *You have those discussions with your colleagues….sat in the office thinking I’m not quite sure about this, it doesn’t sound quite right (Social Worker 2)*.
>
> *I’ve got concerns that these injuries…don’t coincide with what the family are saying (Social Worker 1)*.

However, their professional background and training did not enable them to distinguish between injuries which were likely caused by physical abuse or neglect, and those attributable to other causes, such as accidents:

> *My experience of seeing bruises on adults is very limited….and it’s quite easy to be told by…a carer or a care home that it’s happened this way, but actually…I’m not trained to…say, I’m not sure that’s right or that’s true (Social Worker 6)*.

In this context they were aware there was a risk they could miss or mis-interpret signs and not respond effectively when individuals were at risk or had been injured as a result of physical abuse or neglect. Therefore, they recognised the value of gathering information and opinions from other sources. Prior to the establishment of the Forensic Service, Social Workers reported that they could seek the support of health practitioners, such as GPs and Safeguarding Nurses. However, these practitioners did not themselves have forensic training, and were unable therefore to offer a *forensic slant (Social Worker 5)*, or interpret injuries. Further, these practitioners were not always able to provide timely responses.

In such circumstances, Social Workers appeared to experience uncertainty about how best to proceed when they were concerned that individuals had experienced NAI, and could be left feeling uncertain, uncomfortable and stuck. One Social Worker recalled a situation prior to the introduction of the Forensic Service in which they were suspicious about the causes of an individual’s injuries, but were unsure how to proceed and establish the potential cause. In this case they reported that they were:

> *Really panicking……I didn’t know what to do….literally got left to get on with it and I were like, I don’t even know what I’m doing, having to ask loads of questions and it, it were just out of my comfort zone (Social Worker 1)*.

These findings suggest that Social Workers who are concerned about injuries appear to experience a ‘perfect storm’ in which they are concerned about the welfare of individuals who often cannot provide a clear account of how they were injured; lack the training and knowledge base to interpret injuries and make judgements about the legitimacy of their concerns; and in which health practitioners may also lack the forensic knowledge to offer an opinion and timely guidance. The forensic service seeks to close this gap in respect of knowledge, understanding and practice, and provide informed opinion in respect of injuries, reducing uncertainty.

### Theme 2 – The role of forensic expertise in enhancing knowledge and understanding of non-accidental injury:”It gives us a deeper understanding”

The previous theme identified the challenges for Social Workers (and other practitioners) in recognising injuries which are likely caused by physical abuse or neglect. This theme explores how access to forensic expertise enhanced knowledge and provided important additional information.

The FME delivered training to Social Workers within safeguarding adults teams, helping them to understand *non-accidental injury and what that might look like…and what we should look out for (Social Worker 2)*, as well as the purpose and role of the new service. Joint examination visits attended by Social Workers and the FME provided further opportunities for informal learning. These opportunities for learning and the *exposure….to that other field of professionalism (Social Worker 2)* helped shape and deepen Social Workers’ understanding of non-accidental injury, important signs of which to be aware and take into consideration, and *alters the way that you’re thinking about things and looking at things and resolving cases (Social Worker 4)*.

The new service and the understandings and insights it provided appeared to bolster Social Workers’ professional curiosity, enabling them to ask more questions and request information, while enhancing their confidence that they would be able to use this information:

> *I don’t think that photographs was requested as commonly [previously], purely for the reason, I wouldn’t have known what I was looking for even if we had the photographs, unless it was something really obvious….so I think that it’s also encouraged us to delve deeper and to request more information (Social Worker 4)*.

Social Workers were able to consult the FME when they were concerned about individuals’ injuries and the plausibility of accounts about how these had been sustained, and they valued being able to draw on the expertise of a trained FME in these circumstances. The FME represented a routinely used first point of contact when Social Workers were concerned, unsure or wanted to ‘double check’ or seek advice, although some described also using ‘common sense’ to rule out obvious causes of injury or reviewing case notes for information about how the injury may have been caused. This ability to contact the FME provided rapid access to an expert opinion, and could help inform what further information should be requested and determine whether an examination was indicated.

### Theme 3 –: The role of forensic information in supporting safeguarding decision making and activity: ‘It feels much more robust and much more safe’

Forensic information helped shape decisions about the most likely causes of injury, the level of concern indicated, and the actions which should follow. It represented *a piece of the jigsaw (Social Worker 1)* alongside information and concerns from other sources (such as information from Social Worker examinations of care records and the wider care environment, information on family dynamics), which collectively formed the evidence base upon which safeguarding decisions were made. Forensic information could assist Social Workers to progress beyond feelings of misgiving or doubt, through providing clearer evidence, reducing uncertainty, and leading to *a sounder decision (Social Worker 2):*

> *I think without [FME] being there….we would have never been able to identify why that bruising had occurred in the first place, so it helps us to guide where we need to go next and what we need to be looking at and amending. It would have been really difficult for us, if I’d have gone out on that visit and observed the bruising….I wouldn’t have known and have been able to say whether that was accidental or non-accidental, so it really guided where I went from that point (Social Worker 4)*.
>
> *If we didn’t have that information, we would have been saying potentially it may have been caused by this fall or potentially it might have been this, and there’d be more uncertainty (Social Worker 6)*.

There was relatively little detailed information about the actions that followed on from the examinations. The FME reported that they did not always receive feedback, and Social Workers observed that they often do not hold cases throughout, and so were not always aware of any decisions made or activity undertaken once their involvement ended. However, the findings do suggest that there are different broad categories of injury causation, as outlined below, and that these appear to align with different kinds of safeguarding activity, although further detail is needed to develop this further.

### Medical and accidental causes

In some instances, injuries were attributed to natural causes, such as skin changes associated with ageing which may resemble bruising, or to accidents which appeared unlikely to recur. In one such example, a care home resident was found to have bruising consistent with being gripped, which reportedly occurred when a staff member sought to prevent them falling; this injury was consistent with the account of the incident, which was witnessed. Such interpretations suggested that little or no further action was required, and allowed concerns to be de-escalated.

### Incidents with risk of recurrence

In other cases, the causes of injury were not considered contingent upon abuse or neglect, but causes were identified which suggested that such injuries could recur if no further actions were taken. Examples included unsafe environments which had the potential to cause unintentional injury, or poor practices within the care environment. Once identified, these risks could be mitigated and could inform safety plans and actions intended to reduce the risk of recurrence and to better support the person:

> *We can then maybe put some actions in place to make sure that….the risk is reduced … we’ve put things in place like risk assessments or falls referrals….So even if we get back “no, it’s all fine” we then know that….this needs to be looked at through the care and support and resolve it in a different way than formal inquiry (SW2)*
>
> *Her moving and handling care plan was changed,…. improved her quality of care. So she would have had much better care and obviously not been in pain because…it wouldn’t have resulted in bruising (SW4)*.

### Non-accidental injury

In other cases, the examination findings indicated that the injury had likely been sustained non-accidentally, indicating that further safeguarding investigation and protective actions were required.

The ability to determine likely causes appeared helpful to practitioners, whether or not they suggested that NAI had occurred. Where concerns about abuse and neglect were not substantiated, this appeared to provide reassurance for practitioners and care providers, as well as guidance about the most appropriate responses to prevent ongoing harm:

> *It is sometimes hard to let things rest if you do feel uneasy about it, so knowing that you’ve gone down every avenue possible and looked at it from every way that you can does help (Social Worker 3)*.
>
> *This one that we went out to, potentially…could have been a non-accidental injury but it turned out not to be, so that was also helpful because…[FME] was still able to give that advice on to how to prevent these, this from happening in the future and we could support with that (Social Worker 4)*.

### Overall, the information from the examination helped to guide

*How, why and where we’re going to take our decision making for the next stage or whether we’re quite satisfied that we can close it there and be assured that we know that people are safe (Social Worker 4)*.

Recourse to information from forensic examinations also appeared to strengthen the ‘safeguarding voice’. Social Workers reported being able to better explain to care providers the decisions made and the rationale for these, and to articulate the basis on which funding for enhanced care packages was sought:

> *I was able to evidence why we needed for them to stay in care…I could evidence that absolutely this person could not go back home with the normal care package, if they were to go back home they’d need two carers because of how aggressive the family member was (Social Worker 1)*.
>
> *It also provides information on the enquiry reports, so we can then….share that with providers and say….this is what we’ve found, this is why we’re going to be taking this forward and this is the professional medical opinion that we’ve sought to support that decision (Social Worker 4)*.

However, it was also perceived by Social Workers that the medical voice held greater sway and was considered more authoritative, with other agencies felt to be more likely to listen to an opinion from a clinician and to act on their recommendations:

> *[The FME] was able to give advice and guidance from the medical perspective to the care home and I think, maybe wrong to say it, but coming from a doctor I think often people listen more (Social Worker 3)*.

### Theme 4 – Working together within the new service partnership: ‘It makes us all stronger doesn’t it’

Joint working between Social Workers and the FME when there were concerns about injuries was underpinned by a background urgency and requirement for a rapid and timely response, with examinations ideally needing to take place within 48 hours. Injuries may fade quickly, therefore any visits required should be arranged promptly, and it is important that practitioners can contact one another readily in the context of work which is both unpredictable and requires practitioners to be reactive and responsive.

Social Workers reported that the FME was approachable, accessible and could be easily contacted, responded to calls swiftly, and that joint visits could be arranged readily. However, both Social Workers and the FME were aware of the limitations of a service reliant upon a single practitioner, in which there was no cover during periods of absence. Social Workers recognised the importance of rapid responses when examination visits were required, and reported that they could work flexibly to enable such visits to be prioritised. The FME valued the quality of discussions held with Social Workers, however, observed there could sometimes be difficulties in making contact with social workers and arranging discussions in a timely manner, which they attributed to changed working practices in which social workers may work remotely and spend less time in the office than previously:

> *Very few actually go into the office…..since COVID, we no longer can ring the office up. You can’t ring and speak to a Social Worker in fact very few of them put their phone number on their email (FME)*.

In addition to physical accessibility, accessibility of information was also reported, with Social Workers highlighting the presentation of complex information by the FME in ways which could be understood by practitioners without a medical background. Jargon was avoided and t*here’s no silly question that can be asked which is really good when you’re trying to do an enquiry (Social Worker 5)*.

Social Workers and the FME reported positive working relationships. Underpinning these was an apparent mutual recognition and appreciation of each others’ distinct roles, perspectives, professional skills and abilities, and contribution to adult safeguarding. This involved *knitting together their skills and my skills (FME)*, in which both practitioner groups worked to collect their respective *pieces of the jigsaw, and then try to put them together (FME)*. The importance of working collaboratively and respectfully was highlighted, in which it was important to ensure that the FME and Social Workers could discuss their views and perspectives, and in which there was space for professional challenge and questions, rather than falling back on traditional professional hierarchies:

> *I did feel able to discuss my views….I think it’s important to have both views although I’ve not got medical knowledge I’ve got a lot of experience and knowledge from social care that is important and a lot of experience of going into care homes and looking at different things and being concerned about different things….listening to that gut feeling about something (Social Worker 3)*.
>
> *I think it needs to be an equal relationship. and we can discuss stuff and, if necessary challenge each other professionally about the decisions we’re making (FME)*.

## Discussion

The findings highlight the value of a service which provides access to forensic expertise to support decision making when there are concerns about individuals’ injuries. Social Workers, who often lead on safeguarding enquiries, do not have the training and professional background to equip them to interpret injuries or differentiate between those likely attributable to physical abuse and those more probably arising from other causes. The forensic service appeared to help address this knowledge and training gap.

### The contribution of the Forensic Service to safeguarding decision-making, practice and activity

The findings emphasised that the service had a valuable role in helping ascertain the likely causes of injuries and determining the level of concern warranted. In some instances, injuries appeared to be indicative of underlying medical causes or to have arisen accidentally. In these circumstances, further safeguarding activity was seldom likely to be required. Other injuries which were not considered due to NAI were judged likely to recur if actions were not taken to address the underlying causes (such as risks within the person’s environment). In this context the forensic examination had a valuable role in highlighting activity which was required in order to reduce future risk (and in which, if these actions were not taken, individuals would arguably experience ongoing harm due to neglect and failure to implement changes). Other injuries were considered to be non-accidental and to be caused by abuse and neglect. In these instances further investigation and safeguarding activity to protect the person from ongoing abuse or harm was required. The ability to determine the most appropriate level of concern was helpful in guiding decisions about when cases could appropriately be closed or referred onwards, and helped assure practitioners of the legitimacy of these decisions, while helping allay concerns that important signs had been missed. Conversely, when abuse or neglect were indicated, often stretched resources could be directed towards areas of greatest need. This is of critical importance given the context of contemporary adult safeguarding in which the number and complexity of referrals increases annually (Dixon, 2023; NHS Digital, 2023), and support to prioritise those cases in which further safeguarding investigation and interventions is required can reduce pressure on safeguarding adults teams and ensure proportionate responses.

SARs (Preston-Shoot *et al*., 2020) have highlighted the importance (and sometimes absence) of professional curiosity, which plays an important role in safeguarding and risk assessment, through its emphasis on deep inquiry, respectful challenge, questioning assumptions, encouraging practitioners to ‘think the unthinkable’ and overcome the ‘rule of optimism’ (Burton & Revell, 2017; Thacker *et al*., 2019; Phillips et *al*., 2022). There was some evidence that the forensic project had contributed to such professional curiosity. This included reports that the training Social Workers had received encouraged them to ‘delve deeper’, request further information and question potential causes of harm.

### The importance of multi-disciplinary approaches in adult safeguarding

Although the findings suggest that the inclusion of forensic information has a positive impact on safeguarding practice and decisions when there are concerning injuries, it is also important that this does not take precedence over softer skills which provide important intelligence. Risks of ‘professional deference’ to a practitioner perceived to have higher professional status (such as a GP) have been identified; this may be a barrier to professional curiosity and undermine Social Workers’ confidence to question or challenge their perspectives (Mangan *et al*., 2015; Thacker *et al*., 2019). This was not apparent within the data captured in this study, and Social Workers suggested that recourse to forensic expertise promoted, rather than stifled their professional curiosity. However, they perceived that other people listened to medical practitioners and took their views seriously, highlighting a need for cautious approaches when developing services which foreground medical expertise, to ensure that the perspectives of all practitioners inform decision making. The importance of ensuring that safeguarding decision making does not solely depend on medical opinion is reinforced by the findings of Yonoshiro-Cho *et al*. (2021) who have documented that, while understanding patterns of injury are important, not all individuals who have been physically abused have visible signs of injury, and the absence such signs does not preclude physical abuse. Therefore, while a forensic service can assist in identifying probable causes of injuries and whether they are consistent with abuse or alternative explanations, there may be instances where there are no visible signs. Therefore, there cannot be absolute reliance on medical examination to determine responses, and it is of paramount importance that Social Workers and other practitioners can raise questions and offer alternative viewpoints and evidence, and that forensic and medical opinion is not the sole determinant of safeguarding decisions, based on the presence or absence of injury, and that other concerning signs are attended to.

### Working together

Inter-professional working and relationships have been consistently cited as an important element of adult safeguarding practice (Fyson and Kitson, 2012; Lee *et al*., 2019; Thacker *et al*., 2019). Positive working relationships were reported by both the FME and Social Workers. These reflected aspects of inter-professional working between GPs and Social Workers previously identified as important (Mangan *et al*., 2015). These include trust, respect, flexibility and mutual appreciation of the skills and knowledge each practitioner group brings to safeguarding. In the context of examination of injuries which is time-sensitive work, a further important aspect of inter-professional working was evident in respect of the ease with which practitioners can contact one another. This is necessary to facilitate discussion of cases and arrange visits which are often required at short notice. The move to remote working in response to the COVID-19 pandemic appears to have been retained in some teams in which hybrid working may continue to be undertaken (Pulman and Fenge, 2024). This may lead to challenges for Social Workers, which include isolation from colleagues and peer support, and a loss of team culture and identity (Pulman and Fenge, 2024). The findings from our study identified that these changes to the location of Social Workers also created challenges in contacting Social Workers. Multi-professional safeguarding practice is often reactive, unpredictable and requires ease of contact and swift responses to ensure evidence can be collected. Thus, it is essential that practitioners have reliable and rapid ways of contacting one another, and that, where staff are not in the office, there are effective systems for forwarding messages and facilitating contact.

### Strengths and limitations

This study provides early insights into the contribution of a service which delivers forensic expertise and support to adult safeguarding practice and decision-making. This service is believed to be currently unique within the UK and to date there appear to have been limited similar initiatives internationally. The focus on how the service supports safeguarding Social Workers helps address an evidence gap in respect of forensic services to support adult safeguarding.

There is an asymmetry in the findings in which the strongest voice is that of Social Workers, all of whom endorsed the new service and the contribution it made to practice, focussing their responses on the activity of the FME and giving less detail about their own roles in safeguarding when a forensic practitioner is involved. A more detailed account of the practice of both social workers and FMEs when investigating injuries would help further understanding. While this paper focusses on the professional practice of Social Workers and forensic practitioners there are additional important perspectives to include in future research. The inclusion of those who have been examined and their families and friends (when not implicated in the abuse) represent important voices which can inform understandings of the experience of the examination and the acceptability of subsequent safeguarding decisions and activity. It will be important in future research to consider how to undertake this ethically and emotionally sensitive research, which requires careful consideration to reduce the risk (which can be mitigated but not removed entirely) of further distress and harm for people who have been abused and their families (Sherwood-Johnson and Mackay, 2021), and which take into account the cognitive and communicative difficulties often experienced by those referred to the service. The study would have been strengthened by inclusion of other key agencies, such as the Police and care providers.

## Conclusion

This study presents findings drawn from data captured during the first year of delivery of a forensic service to support adult safeguarding, from the perspective of safeguarding Social Workers and the FME who conducted examinations and interpretation of injuries. The development of the service recognises and responds to the challenges of ascertaining whether individuals have been abused or neglected and brings a further significant perspective to adult safeguarding in cases of suspected or possible physical abuse or neglect. The findings indicate that the service brought new understanding and skills to the often-uncertain landscape of adult safeguarding, and was considered to assist safeguarding decision-making and confidence in the robustness of these decisions. A valuable contribution of the Forensic Service was its ability to help differentiate between injuries which appeared due to accidental causes or underlying medical conditions, those which were likely to recur, and those which were most probably due to abuse and neglect. This contributed to providing the most appropriate responses, and appeared to help ensure that safeguarding activity is proportionate, in an area of practice in which there are ongoing increases in referral and demand.

## Data Availability

Due to the sensitivity of the data this is not available to other researchers.

